# IMPACT OF THE COVID-19 PANDEMIC ON ROUTINE HIV CARE AND ANTIRETROVIRAL TREATMENT OUTCOMES IN KENYA: A NATIONALLY REPRESENTATIVE ANALYSIS

**DOI:** 10.1101/2023.09.04.23294973

**Authors:** Davies O. Kimanga, Valeria N.B. Makory, Amin S. Hassan, Faith Ngari, Margaret M. Ndisha, Kennedy J. Muthoka, Lydia Odero, Gonza O. Omoro, Appolonia Aoko, Lucy Ng’ang’a

## Abstract

**Background:** The COVID-19 pandemic adversely disrupted global health service delivery. We aimed to assess impact of the pandemic on same-day HIV diagnosis/ART initiation, six-months non-retention and initial virologic non-suppression (VnS) among individuals starting antiretroviral therapy (ART) in Kenya.

**Methods:** Individual-level longitudinal service delivery data were analysed. Random sampling of individuals aged >15 years starting ART between April 2018 – March 2021 was done. Date of ART initiation was stratified into pre-COVID-19 (April 2018 – March 2019 and April 2019 – March 2020) and COVID-19 (April 2020 – March 2021) periods. Mixed effects generalised linear, survival and logistic regression models were used to determine the effect of COVID-19 pandemic on same-day HIV diagnosis/ART initiation, six-months non-retention and VnS, respectively.

**Results:** Of 7,046 individuals sampled, 35.5%, 36.0% and 28.4% started ART during April 2018 – March 2019, April 2019 – March 2020 and April 2020 – March 2021, respectively. Compared to the pre-COVID-19 period, the COVID-19 period had higher same-day HIV diagnosis/ART initiation (adjusted risk ratio [95% CI]: 1.09 [1.04–1.13], p<0.001) and lower six-months non-retention (adjusted hazard ratio [95% CI]: 0.66 [0.58–0.74], p<0.001). Of those sampled, 3,296 (46.8%) had a viral load test done at a median 6.2 (IQR, 5.3–7.3) months after ART initiation. Compared to the pre-COVID-19 period, there was no significant difference in VnS during the COVID-19 period (adjusted odds ratio [95% CI]: 0.79 [95%% CI: 0.52–1.20], p=0.264).

**Conclusions:** In the short term, the COVID-19 pandemic did not have an adverse impact on HIV care and treatment outcomes in Kenya. Timely, strategic and sustained COVID-19 response may have played a critical role in mitigating adverse effects of the pandemic and point towards maturity, versatility and resilience of the HIV program in Kenya. Continued monitoring to assess long-term impact of the COVID-19 pandemic on HIV care and treatment program in Kenya is warranted.

## BACKGROUND

In January 2020, the World Health Organization (WHO) reported emergence of the novel severe acute respiratory coronavirus 2 (SARS-CoV-2), the causative agent for COVID-19 [1]. By the end of 2021, WHO estimated a global excess mortality of 14.9 million, representing 9.5 million more fatalities than those directly attributable to COVID-19 [2]. At onset of the pandemic and in the absence of efficacious prophylactic or therapeutic interventions, governments worldwide re-purposed already limited resources to enforce non-pharmaceutical interventions to limit spread. These included social-distancing, face-masking, handwashing, closure of learning institutions and places of worship, travel restrictions, quarantine for exposed, isolation for confirmed infections, curfews and partial or complete lockdowns. Whilst these measures had a positive effect on mitigating the spread of COVID-19 [3–6], they may have adversely disrupted health service delivery, including HIV care and treatment programs.

Disruptions in routine HIV care and treatment program may be disentangled into three domains: (i) disruption in supply-chain of commodities, either directly from enforcement of travel restrictions, or indirectly from re-purposing of fiscal and infrastructural resources for COVID-19 emergency response; (ii) disruptions in the workforce, either directly from deployments to COVID-19 related services and from avoidance of workplace due to fear of nosocomial SARS-CoV-2 acquisition, or indirectly from mandatory quarantine or isolation of frontline health care workers; and (iii) disruption in health-seeking behaviour, either directly from hesitancy to seek services from health facilities (perceived as hotspots for SARS-CoV-2 infections), or indirectly from inaccessibility of health facilities due to travel restrictions and lockdowns. Combined, these disruptions may have negatively impacted HIV care and treatment service delivery, including HIV testing and diagnosis, early linkage to care and continuity in supply of life-saving antiretroviral therapy (ART).

Early ART is associated with reduced HIV-related morbidity and mortality [7–10], rapid and sustained virologic suppression [8, 9, 11–13] and reduced risk of onward HIV transmission [14–16]. Benefits conferred by early ART motivated development of WHO guidelines recommending immediate ART to all HIV infected individuals regardless of clinical, immunological or virological status, commonly referred as the test-and-treat policy [17]. We recently demonstrated that same-day HIV diagnosis/ART initiation increased from 15% in 2015 to 52% in 2018 [18], suggesting significant strides in scale up of the test-and-treat policy in Kenya. COVID-19 related disruptions threaten to reverse these gains, though its impact on time from HIV diagnosis to ART initiation in Kenya remains unknown.

Retention in the HIV care and treatment continuum is also critical towards achieving population-level virologic suppression. In a systematic review of 123 publications published between 2008 and 2013 from low-and-middle-income countries (LMIC), retention at 12, 24 and 36 months after ART initiation was estimated at 78%, 71% and 69% respectively [19]. Early treatment interruptions not only result in selection of HIV drug resistance mutations [20], but also pose a threat for onward HIV transmission [21]. COVID-19 related disruptions may have negatively impacted early retention in Kenya, though this is not yet documented.

Emphasis on early ART and retention is aimed at attaining rapid and sustained virologic suppression. The UNAIDS has set ambitious 95-95-95 targets towards ending the HIV epidemic by 2030, with the last target aimed at achieving 95% virologic suppression amongst ART-experienced individuals [22]. In a systematic review of 49 studies from LMIC, virologic suppression after twelve months of ART was estimated at 84% [23]. In Kenya, data from population-based surveys suggest an increase in virologic suppression amongst ART-experienced adults, from an estimated 39% in 2012 to 91% in 2018 [24, 25]. These estimates suggest Kenya is well on track towards achieving the UNAIDS targets on virologic suppression. However, COVID-19 related disruptions threaten to veer the country off the track towards attaining epidemic control.

Unintended disruptions from the COVID-19 pandemic threaten to erode gains made in the fight against the HIV epidemic. We aimed to assess impact of the COVID-19 pandemic on time from HIV diagnosis to ART initiation, six months non-retention and initial virologic non-suppression amongst individuals starting ART in Kenya.

## METHODS

### Study setting

In Kenya, the first case of SARS-CoV-2 infection was reported on March 12^th^, 2020 [26]. By the end of March 2021, the country had reported >130,000 confirmed cases and >2,000 COVID-19 associated fatalities [27]. During this time, the government enforced nation-wide non-pharmaceutical interventions including social distancing, face masking, hand washing, quarantine for the exposed, isolation for confirmed infections and dusk-to-dawn curfews. Initially, closure of learning institutions, restaurants, bars, religious places of worship and ban on international travel were also imposed [28]. COVID-19 vaccinations started on March 08^th^, 2021, initially targeting health workers, teachers, and security personnel. Over time, the elderly, followed by the general adult population were eligible. By end of March 2021, Kenya had experienced two waves of the pandemic and was in the middle of the third wave. The two waves lasted between June to August 2020 and October to December 2020 [27]. The waves affected counties differentially. The first wave adversely affected Nairobi, Mandera and Coastal Kenya counties including Kilifi, Mombasa and Kwale. The second wave adversely affected Nairobi and neighboring counties including Kiambu, Nakuru, Machakos and Kajiado. Affected counties were declared SARS-CoV-2 high infection zones (HIZ) (Figure 1 [a]). Additional interventions including stricter enforcement of COVID-19 protocols, longer curfew hours and partial lockdowns (restriction of movement into and out of HIZs) were imposed in HIZ. The first dose of the COVID-19 vaccine was introduced in March 2021.

**Figure 1.**
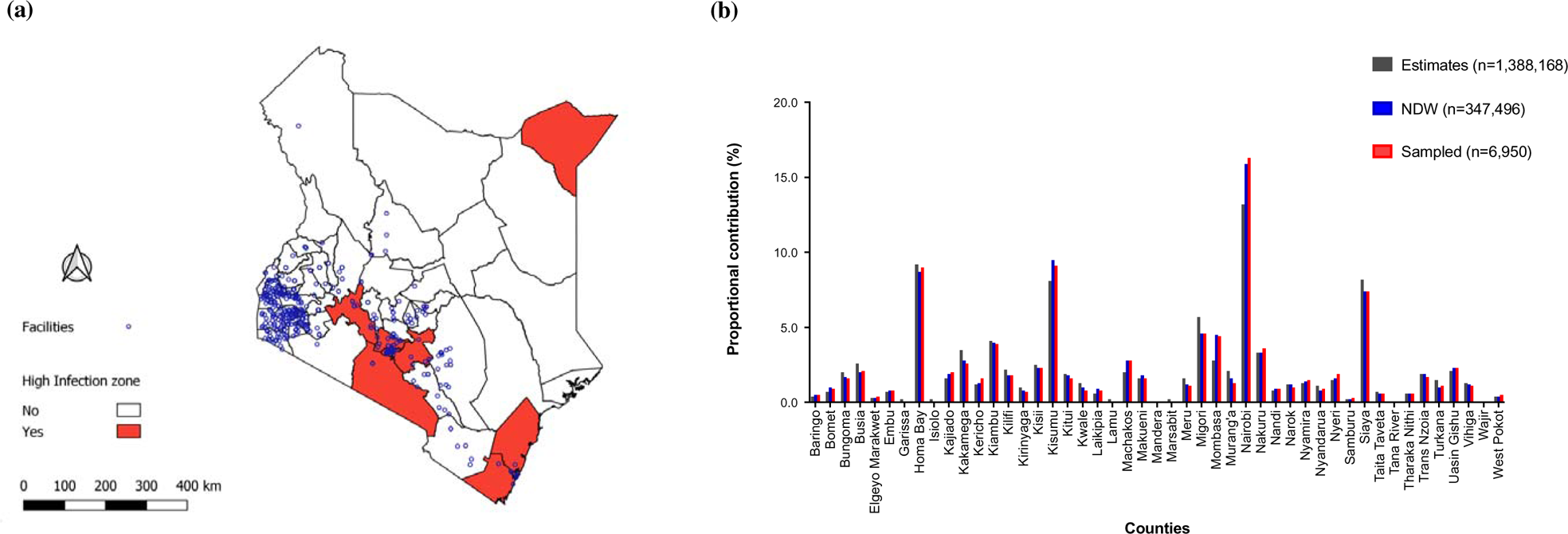
(a) Map showing the distribution of health facilities transmitting electronic medical records (EMR) data to the National Data Warehouse (NDW). Polygons represent the 47 counties (regional administrative units), with red colored polygons representing the high infection zone (HIZ) counties. (b) Graph showing the distribution of estimated number of people living with HIV (PLWH) in Kenya (n = 1,388,168), number of PLWH starting combination antiretroviral therapy between April 2018 and March 2021 in the NDW (n = 352,322), and the number of PLWH randomly sampled and included in the analysis (n = 7,046).

### Study design

Longitudinal data archived at the national data warehouse (NDW) were analyzed. The NDW is a centralized repository of individual-level routine HIV program data transmitted from electronic medical records (EMR) deployed in health facilities offering HIV care and treatment services in Kenya. Routinely collected HIV service delivery data are extracted, checked for duplicate entries, de-identified and electronically transmitted to the NDW. By end of 2020, the NDW hosted data from ∼2 million individuals ever started on ART from ∼1,500 health facilities covering 44 of the 47 counties in Kenya (Figure 1). The three counties that have never contributed data to the repository are from the historically marginalized North Eastern region of Kenya and were all estimated to have zero new HIV infections in 2018 [29].

### Eligibility criteria and sampling strategy

The sampling framework comprised individuals age >15 years starting ART during April 2018 – March 2021. Volunteers starting ART after March 2021 were not considered to avoid potential confounding that may have resulted from introduction of the COVID-19 vaccines on analysis end-points. Two percent of the sampling framework was randomly sampled. A *post-hoc* sample size calculation was done. We assumed 52%, 78% and 90% of the randomized population had same-day HIV diagnosis/ART initiation, were retained in care six months after ART initiation and achieved virologic suppression within 12 months of ART initiation, respectively [18, 19, 25]. We also assumed one-third (33%) of the randomized population started ART during the COVID-19 period, defined as the period between April 20 – March 21. Based on these assumptions, the sampled population conferred 88%, 96% and 99% power to detect a conservative 4% relative difference in same-day HIV diagnosis/ART initiation, six-months retention and virologic suppression, respectively, between the pre-COVID-19 and the COVID-19-periods (two-sided alpha, 0.05). Further, we assumed 33% of the randomized population were from the nine counties that were declared HIZ [30]. Additional restrictions imposed on these counties were considered *a-priori* as effect modifying. Results were thus further stratified by whether individuals were from within or outside the HIZ. The sampled population from within the HIZ (the smaller stratum) conferred 76%, 90% and 99% power to detect a modest 6% relative difference in same-day HIV diagnosis/ART initiation, six-months retention and virologic suppression, respectively, between the pre-COVID-19 and the COVID-19 periods (two-sided alpha, 0.05).

### Definition of indicators

Calendar periods were defined according to when individuals started ART as follows: pre-COVID-19 period (April 01st 2018 – March 31st 2019, and April 01st 2019 – March 31st 2020) and the COVID-19 period (April 01st 2020 – March 31st 2021). End-points included; (i) time from a HIV diagnosis to ART initiation, defined as same-day HIV diagnosis/ART initiation, (ii) short term non-retention, defined as individuals who were either dead or lost to follow up (LTFU, missed scheduled appointments plus three months grace period) within six months of ART initiation, and (iii) initial virologic non-suppression (VnS), based on the first viral load test done within 12 months of ART initiation and defined as HIV RNA >1000 copies/ml.

### Data analysis

Same-day HIV diagnosis/ART initiation is not a rare (>50%) endpoint. Thus, univariable and multivariable generalized linear models (glm) were used to determine effect of the COVID-19 period on same-day HIV diagnosis/ART initiation. Crude and adjusted risk ratios, 95% confidence intervals (CIs) and p-values were reported.

Time-to-event analyses were used to determine time from ART initiation to non-retention over a six-months follow-up period. Because some individuals started ART during the pre-COVID-19 period, but their follow-up crossed over into the COVID-19 period, Lexis expansion was applied and follow-up time split accordingly, either to pre-COVID-19 or COVID-19 periods [31]. Univariable and multivariable survival regression models were used to determine effect of COVID-19 period on non-retention. Crude and adjusted hazard ratios (HR), 95% CIs and p-values were reported.

Initial VnS may be considered a rare (<10%) endpoint. Thus, univariable and multivariable logistic regression models were used to determine effect of the COVID-19 period on initial VnS. Crude and adjusted odd ratios, 95% CIs and p-values were reported.

In all regression models, variables with a p-value <0.05 from the univariate analysis were carried forward to the multivariate models. Hierarchical mixed effect modeling was applied to control for within-and-between counties variations. All analyses were done using Stata I/C (version 15.1).

### Ethical considerations

This retrospective analysis was part of a National HIV program evaluation exercise. Ethics approval to waive need for informed consenting was obtained from the Africa Medical Research Foundation (AMREF) Ethics Scientific Review Committee, Kenya (AMREF-ESRC P716/2019). The analysis was reviewed in accordance with the U.S. Centers for Disease Control and Prevention (CDC) human research protection procedures and was determined to not meet the definition of research as defined in 45CFR46.102. All the data used in the analysis were fully anonymized. Thus, authors did not have access to information that could identify individual participants during or after data collection.

## RESULTS

### Characteristics of participants

By end of March 2021, the NDW hosted individual-level data from ∼2 million individuals ever started on ART. Of these, 352,322 were aged >15 years and started ART during April 2018 – March 2021 (Figure 2).

**Figure 2.**
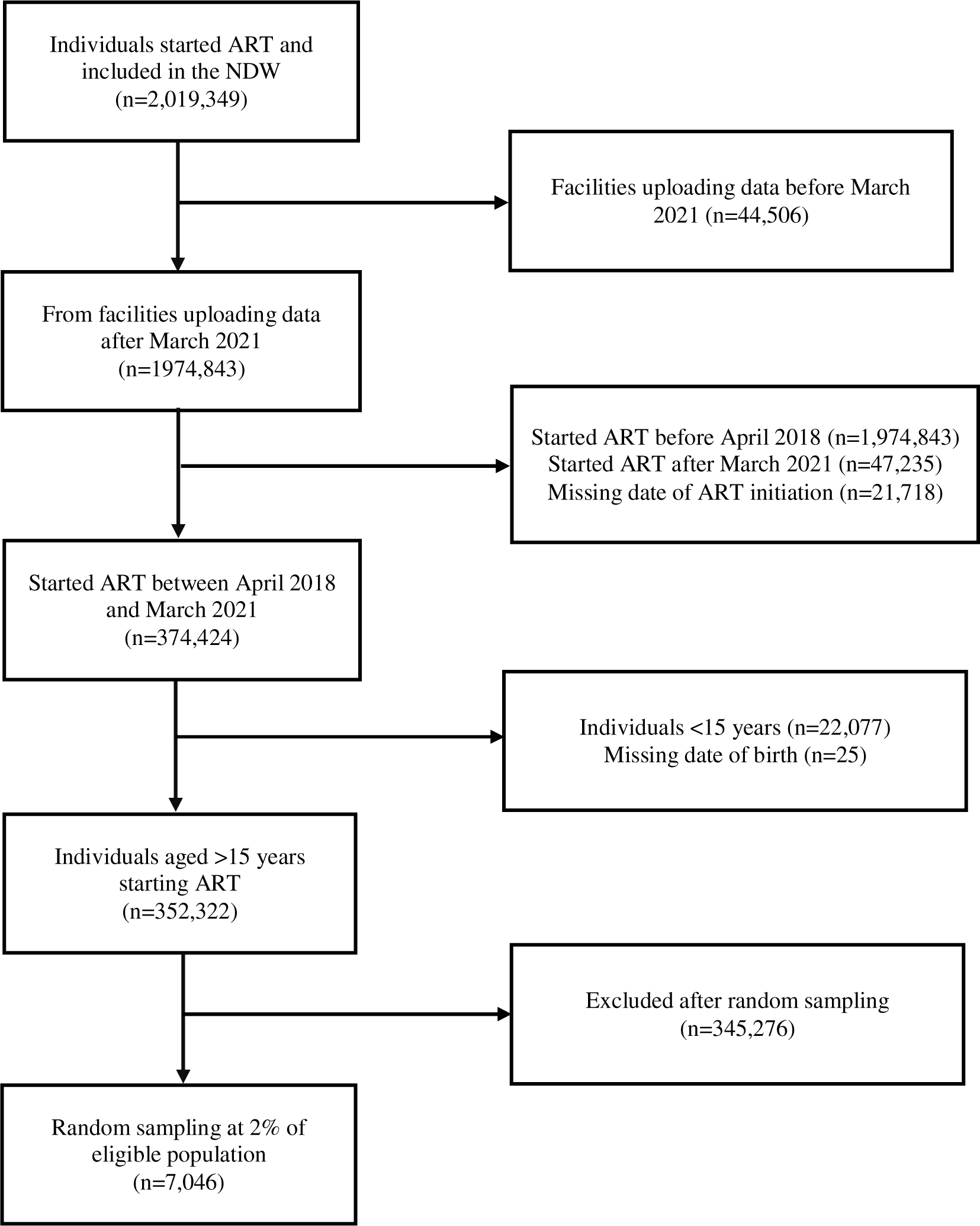
Flow chart showing distribution of HIV infected individuals starting combination antiretroviral therapy (ART) included in the national data warehouse (NDW) sampling framework and random selection of individuals included in the analysis.

Of these, 7046 (2.0%) were randomly sampled. The proportional distribution of sampled individuals by counties was consistent with that from the NDW sampling frame and the most recent national HIV estimates report of 2018 (Figure 1 [b]). Compared to those that were not sampled, there were no differences in the characteristics of sampled individuals (Table 1).

**Table 1.**
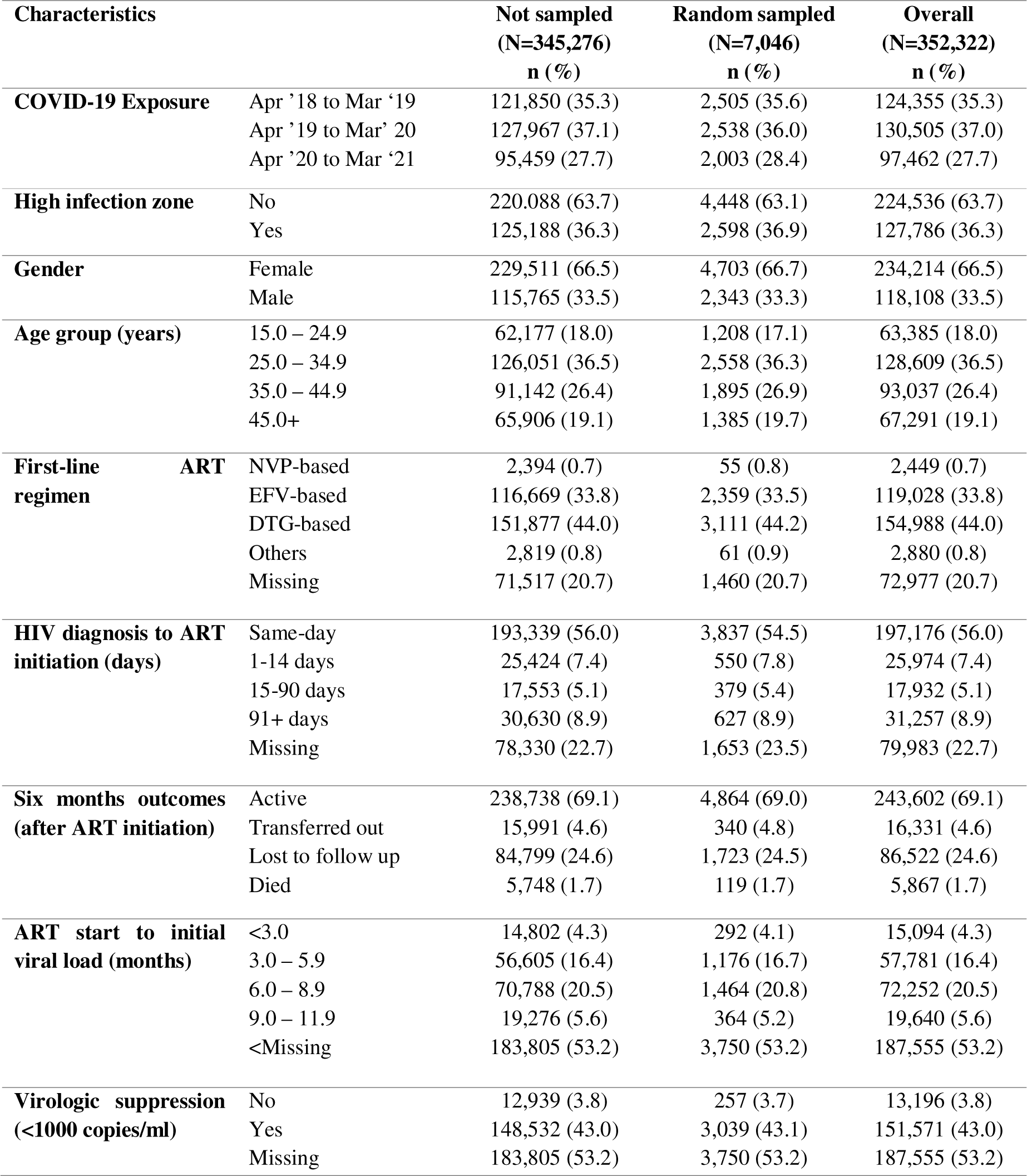
A comparison of the general eligible and randomly sampled population of HIV infected individuals aged >15 years at the start of combination antiretroviral therapy using data from the national data warehouse (April 01^st^ 2018 to March 31^st^ 2021, N=352,322).

Of those sampled, 2,505 (35.5%), 2,538 (36.0%) and 2,003 (28.4%) started ART during April 2018 – March 2019, April 2019 – March 2020 and April 2020 – March 2021, respectively. The majority were female (n=4,703 [66.7%]). The proportion of individuals started on a dolutegravir (DTG-based) regimen increased from 14.6% during April 2018 – March 2019 to 82.8% during April 2020 – March 2021 (Table S1). Overall, 2,598 (36.9%) were from the HIZ. Compared to individuals from outside the HIZ, there were no major differences in the characteristics of those from the HIZ (Table 2).

**Table 2.** Distribution of HIV infected individuals >15 years old, starting combination antiretroviral therapy, included in the national data warehouse sampling framework and randomly sampled, by COVID-19 exposure status and high infection zones in Kenya (April 01^st^ 2018 to March 31^st^ 2021, N=7,046).

| Characteristics |  | Pre-COVID-19 period |  |  |  | COVID-19 period |  | Overall |  |
| --- | --- | --- | --- | --- | --- | --- | --- | --- | --- |
|  |  | Apr '18 to Mar '19 |  | Apr '19 to Mar '20 |  | Apr '20 to Mar '21 |  | HIZ, No<br>(N=4448)<br>n (%) | HIZ, Yes<br>(Yes, N=2598)<br>n (%) |
|  |  | HIZ, No<br>(N=1605)<br>n (%) | HIZ, Yes<br>(N=900)<br>n (%) | HIZ, No<br>(N=1582)<br>n (%) | HIZ, Yes<br>(N=956)<br>n (%) | HIZ, No<br>(N=1261)<br>n (%) | HIZ, Yes<br>(N=742)<br>n (%) |  |  |
| **Gender** | Female | 1069 (66.6) | 612 (68.0) | 1039 (65.7) | 653 (68.3) | 848 (67.3) | 482 (65.0) | 1956 (66.5) | 1747 (67.2) |
|  | Male | 536 (33.4) | 288 (32.0) | 543 (34.3) | 303 (31.7) | 413 (32.7) | 260 (35.0) | 1492 (33.5) | 851 (32.8) |
| **Age group**<br>**(years)** | 15.0 – 24.9 | 304 (18.9) | 126 (14.0) | 279 (17.6) | 155 (16.2) | 230 (18.2) | 114 (15.4) | 813 (18.3) | 395 (15.2) |
|  | 25.0 – 34.9 | 589 (36.7) | 320 (35.6) | 566 (35.8) | 365 (38.2) | 459 (36.4) | 259 (34.9) | 1614 (36.3) | 944 (36.3) |
|  | 35.0 – 44.9 | 386 (24.1) | 266 (29.6) | 423 (26.7) | 243 (25.4) | 370 (29.3) | 207 (27.9) | 1179 (26.5) | 716 (27.5) |
|  | 45.0+ | 326 (20.3) | 188 (20.9) | 314 (19.9) | 193 (20.2) | 202 (16.0) | 162 (21.8) | 842 (18.9) | 543 (20.9) |
| **First-line ART**<br>**regimen** | NVP-based | 27 (1.7) | 24 (2.7) | 1 (0.1) | 2 (0.2) | 1 (0.1) | 0 (0.0) | 29 (0.7) | 26 (1.0) |
|  | EFV-based | 857 (53.4) | 518 (57.6) | 533 (33.7) | 330 (34.5) | 75 (6.0) | 46 (6.2) | 1,465 (32.9) | 894 (34.4) |
|  | DTG-based | 248 (15.5) | 117 (13.0) | 690 (43.6) | 398 (41.6) | 1038 (82.3) | 620 (83.6) | 1,976 (44.4) | 1,135 (43.7) |
|  | Others | 12 (0.8) | 22 (2.4) | 10 (0.6) | 10 (1.1) | 4 (0.3) | 3 (0.4) | 26 (0.6) | 35 (1.4) |
|  | Missing | 461 (28.7) | 219 (24.3) | 348 (22.0) | 216 (22.6) | 143 (11.3) | 73 (9.8) | 952 (21.4) | 508 (19.6) |
| **HIV diagnosis**<br>**to ART**<br>**initiation**<br>**(days)** | Same-day | 915 (57.0) | 320 (35.6) | 897 (56.7) | 411 (43.0) | 870 (69.0) | 424 (57.1) | 2,682 (60.3) | 1,155 (44.4) |
|  | 1-14 days | 146 (9.1) | 66 (7.3) | 141 (8.9) | 48 (5.0) | 103 (8.2) | 46 (6.2) | 390 (8.8) | 160 (6.2) |
|  | 15-90 days | 99 (6.2) | 66 (7.3) | 82 (5.2) | 46 (4.8) | 46 (3.7) | 40 (5.4) | 227 (5.1) | 152 (5.9) |
|  | 91+ days | 183 (11.4) | 150 (16.7) | 111 (7.0) | 86 (9.0) | 57 (4.5) | 40 (5.4) | 351 (7.9) | 276 (10.6) |
|  | Missing | 262 (16.3) | 298 (33.1) | 351 (22.2) | 365 (38.2) | 185 (14.7) | 192 (25.9) | 798 (17.9) | 855 (32.9) |
| **ART start to**<br>**initial viral**<br>**load (months)** | <3.0 | 69 (4.3) | 52 (5.8) | 54 (3.4) | 46 (4.8) | 40 (3.2) | 31 (4.2) | 163 (3.7) | 129 (5.0) |
|  | 3.0 – 5.9 | 306 (19.1) | 153 (17.0) | 276 (17.5) | 188 (19.7) | 161 (12.8) | 92 (12.4) | 743 (16.7) | 433 (16.7) |
|  | 6.0 – 8.9 | 367 (22.9) | 222 (24.7) | 419 (26.5) | 216 (22.6) | 161 (12.8) | 79 (10.7) | 947 (21.3) | 517 (19.9) |
|  | 9.0 – 11.9 | 103 (6.4) | 41 (4.6) | 123 (7.8) | 64 (6.7) | 23 (1.8) | 10 (1.4) | 249 (5.6) | 115 (4.4) |
|  | <Missing | 760 (47.4) | 432 (48.0) | 710 (44.9) | 442 (46.2) | 876 (69.5) | 530 (71.4) | 2,346 (52.7) | 1,404 (54.0) |

### Effect of the COVID-19 pandemic on same-day HIV diagnosis and ART initiation

Of those sampled, 5,393 (76.5%) had a date of HIV diagnosis and contributed data on time to ART initiation. Overall, same-day HIV diagnosis/ART initiation increased from 63.5% during April 2018 - March 2019 to 79.6% during April 2020-March 2021 (Figure 3 [a]). Same-day HIV diagnosis/ART initiation was significantly higher during the COVID-19 period compared to the pre-COVID-19 period (adjusted risk ratio, aRR [95% CI], p-value: 1.09 [1.04–1.13], p<0.001) (Table S2).

**Figure 3.**
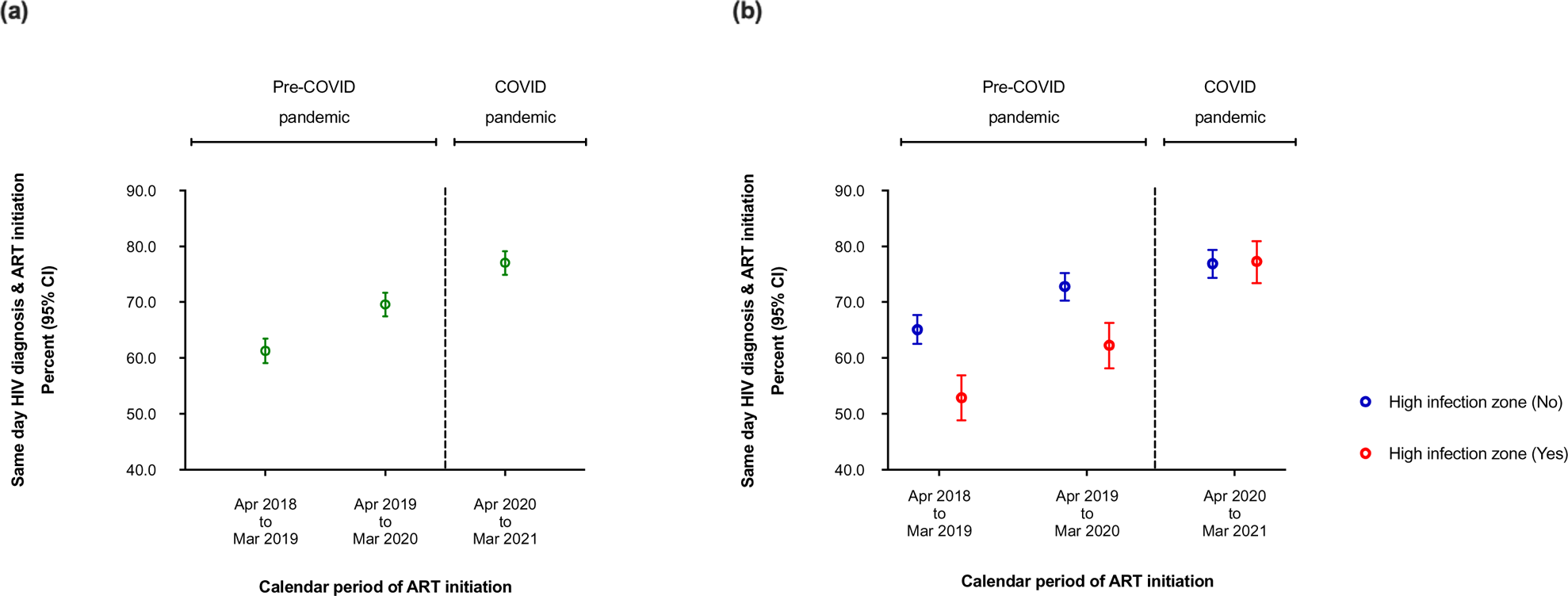
(a) Graph showing time from HIV diagnosis to ART initiation by COVID-19 exposure calendar period when compared to the period prior to the pandemic, and (b) by high infection zone counties, amongst HIV infected individuals >15 years using data from the national data warehouse sampling framework in Kenya (April 01^st^ 2018 to March 31^st^ 2021, N=7,046).

From the HIZ, same-day HIV diagnosis/ART initiation increased from 53.2% during April 2018 – March 2019 to 77.1% during April 2020 – March 2021, while that from outside the HIZ increased from 68.1% to 80.9% during the same period (Figure 3 [b]). From outside the HIZ, same-day HIV diagnosis/ART initiation was significantly higher during the COVID-19 period compared to the pre-COVID-19 period (1.09 [1.04–1.14], p<0.001). Similarly, and from within the HIZ, same-day HIV diagnosis/ART initiation was significantly higher during the COVID-19 period compared to the pre-COVID-19 period (1.10 [1.01–1.19], p=0.020) (Table 3).

**Table 3.** Effect of the COVID-19 pandemic, defined as the period after the first documented case when compared to the period prior to the pandemic, on time from a HIV diagnosis to combination antiretroviral therapy start (Same-day HIV diagnosis and ART initiation) amongst HIV infected individuals aged >15 years using data from the national data warehouse sampling framework in Kenya (April 01^st^ 2018 to March 31^st^ 2021, N=7,046)*.

| Characteristics |  | High infection zone (No, n=3,650) |  |  |  | High infection zone (Yes, n=1,743) |  |  |  |
| --- | --- | --- | --- | --- | --- | --- | --- | --- | --- |
|  |  | Crude RR<br>(95% CI) | p-value | Adjusted RR<br>(95% CI) | p-value | Crude RR<br>(95% CI) | p-value | Adjusted RR<br>(95% CI) | p-value |
| **Pandemic periods** | Pre-COVID-19 | Ref |  | Ref |  | Ref |  | Ref |  |
|  | COVID-19 | 1.13 (1.09 – 1.18) | <0.001 | 1.09 (1.04 – 1.14) | <0.001 | 1.22 (1.14 – 1.30) | <0.001 | 1.10 (1.01 – 1.19) | 0.020 |
| **Gender** | Female | Ref |  |  |  | Ref |  |  |  |
|  | Male | 0.97 (0.93 – 1.01) | 0.146 | - |  | 0.96 (0.89 – 1.02) | 0.243 | - |  |
| **Age group (years)** | 15.0 – 24.9 | 1.21 (1.13 – 1.29) |  | 1.18 (1.11 – 1.26) |  | 1.13 (1.02 – 1.26) |  | 1.16 (1.03 – 1.28) |  |
|  | 25.0 – 34.9 | 1.12 (1.05 – 1.19) |  | 1.10 (1.04 – 1.17) |  | 1.01 (0.98 – 1.12) |  | 1.10 (1.00 – 1.20) |  |
|  | 35.0 – 44.9 | 1.11 (1.04 – 1.18) |  | 1.10 (1.03 – 1.16) |  | 0.94 (0.85 – 1.05) |  | 0.97 (0.88 – 1.08) |  |
|  | 45.0+ | Ref | <0.001 | Ref | <0.001 | Ref | 0.002 | Ref |  |
|  |  |  |  |  |  |  |  |  | 0.001 |
| **First-line ART regimen** | EFV-based | Ref |  | Ref |  | Ref |  | Ref |  |
|  | DTG-based | 1.04 (1.00 – 1.08) |  | 1.02 (0.97 – 1.07) |  | 1.22 (1.13 – 1.32) |  | 1.17 (1.06 – 1.28) |  |
|  | Others | 0.17 (0.05 – 0.52) |  | 0.17 (0.06 – 0.53) |  | 0.22 (0.04 – 1.36) |  | 0.22 (0.03 – 1.42) |  |
|  | Missing | 0.89 (0.85 – 0.95) | <0.001 | 0.90 (0.86 – 0.96) | <0.001 | 0.97 (0.88 – 1.08) | <0.001 | 0.97 (0.88 – 1.07) | <0.001 |

\*Missing date of HIV diagnosis amongst individuals from infection zone (no, n=798 [17.9%]) and (yes, n=855 [32.9%]).

### Effect of the COVID-19 pandemic on six months non-retention

Of those sampled, 4,864 (69.0%), 340 (4.8%), 1,723 (24.4%) and 119 (1.7%) were actively on follow up, transferred, LTFU or reported dead, respectively, six months after ART initiation. Overall, non-retention (n=1,842 [26.1%]) reduced from 10.0/100 person-months-observations (pmo) during April 2018 – March 2019 to 4.3/100 pmo during April 2020 – March 2021 (Figure 4 [a]). When compared to the pre-COVID-19 period, the COVID-19 period had a significantly lower non-retention rate (adjusted hazard ratio, aHR [95% CI], p-value: 0.66 [0.58–0.74], p<0.001) (Table S3).

**Figure 4.**
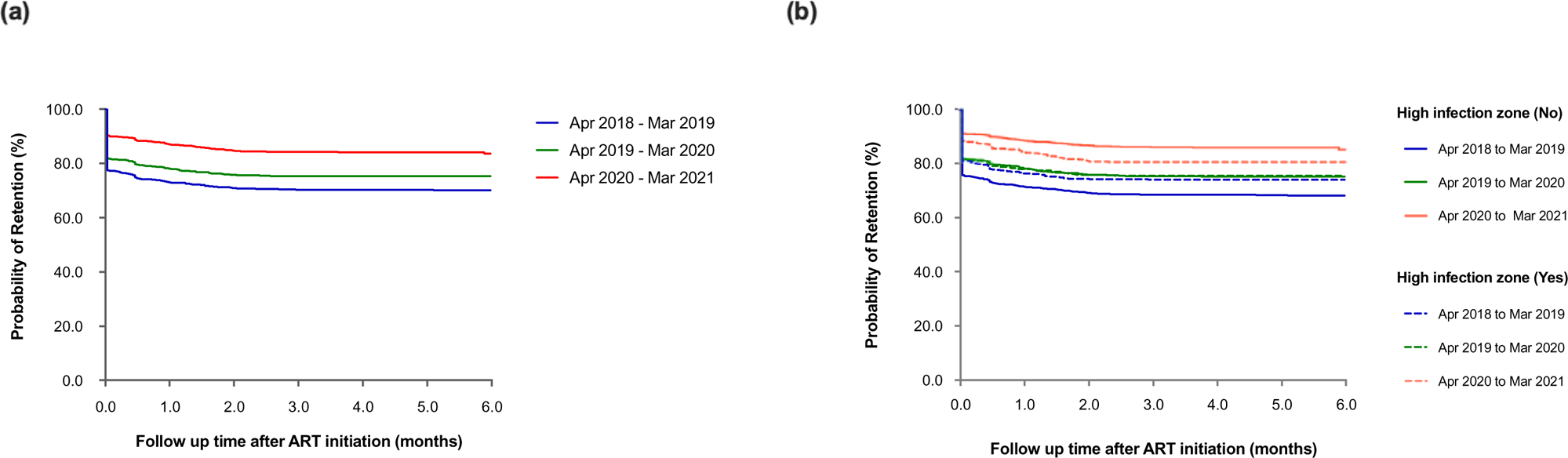
(a) Graph showing six-months retention after ART initiation by COVID-19 exposure calendar period when compared to the period prior to the pandemic, and (b) by high infection zone counties, amongst HIV infected individuals >15 years using data from the national data warehouse sampling framework in Kenya (April 01^st^ 2018 to March 31^st^ 2021, N=7,046).

Non-retention from the HIZ reduced from 9.9/100 pmo during April 2018 – March 2019 to 6.4/100 pmo during April 2020 – March 2021, while that from outside the HIZ reduced from 10.1/100 pmo to

4.4/100 pmo during the same period (Figure 4 [b]). From outside the HIZ, non-retention during the COVID-19 period was significantly lower, compared to the pre-COVID-19 period (0.64 [95% CI: 0.57–0.73], p<0.001). From the HIZ, there was no significant difference in non-retention during the COVID-19 pandemic, compared to the pre-COVID-19 period (0.88 [95% CI: 0.72-1.07], p=0.196) (Table 4).

**Table 4.** Effect of the SARS-CoV-2 pandemic, defined as defined as the period after the first documented case when compared to the period prior to the pandemic, on attrition from antiretroviral therapy care amongst HIV infected individuals >15 years using data from the national data warehouse sampling framework in Kenya (April 01^st^ 2018 to March 31^st^ 2021, N=7,046).

| Characteristics |  | High infection zone (No, n=4,484) |  |  |  | High infection zone (Yes, n=2,466) |  |  |  |
| --- | --- | --- | --- | --- | --- | --- | --- | --- | --- |
|  |  | Crude HR<br>(95% CI) | p-value | Adjusted HR<br>(95% CI) | p-value | Crude HR<br>(95% CI) | p-value | Adjusted HR<br>(95% CI) | p-value |
| **Pandemic periods** | Pre-COVID-19 | Ref |  | Ref |  | Ref |  | Ref |  |
|  | COVID-19 | 0.47 (0.40 – 0.54) | <0.001 | 0.64 (0.57 – 0.73) | <0.001 | 0.70 (0.59 – 0.83) | <0.001 | 0.88 (0.72 – 1.07) | 0.196 |
| **Gender** | Female | Ref |  |  |  | Ref |  | Ref |  |
|  | Male | 1.05 (0.93 – 1.19) | 0.425 | - | - | 1.25 (1.07 – 1.46) | 0.005 | 1.37 (1.16 – 1.63) | <0.001 |
| **Age group (years)** | 15.0 – 24.9 | 1.19 (0.99 – 1.45) |  |  |  | 1.20 (0.93 – 1.54) |  |  |  |
|  | 25.0 – 34.9 | 1.11 (0.94 – 1.31) |  |  |  | 1.12 (0.91 – 1.38) |  |  |  |
|  | 35.0 – 44.9 | 1.09 (0.92 – 1.30) |  |  |  | 1.01 (0.81 – 1.27) |  |  |  |
|  | 45.0+ | Ref | 0.339 | - | - | Ref | 0.412 | - | - |
| **First-line ART regimen** | EFV-based | Ref |  | Ref |  | Ref |  | Ref |  |
|  | DTG-based | 0.70 (0.60 – 0.80) |  | 0.89 (0.78 – 1.00) |  | 0.76 (0.64 – 0.92) |  | 0.72 (0.57 – 0.89) |  |
|  | Others | 0.54 (0.28 – 1.05) |  | 1.15 (0.81 – 1.62) |  | 1.81 (1.19 – 2.74) |  | 1.62 (1.06 – 2.46) |  |
|  | Missing | 2.08 (1.80 – 2.40) | <0.001 | 1.98 (1.77 – 2.23) | <0.001 | 1.63 (1.34 – 1.98) | <0.001 | 1.60 (1.31 – 1.96) | <0.001 |
| **Same-day HIV diagnosis and ART start** | No | Ref |  |  |  | Ref |  | Ref |  |
|  | Yes | 1.15 (1.00 – 1.34) |  |  |  | 1.19 (0.97 – 1.47) |  | 1.37 (1.11 – 1.69) |  |
|  | Missing | 1.16 (0.95 – 1.41) | 0.159 | - | - | 1.53 (1.23 – 1.90) | <0.001 | 1.64 (1.32 – 2.04) | <0.001 |

### Effect of the COVID-19 pandemic on initial virologic non-suppression

Of those sampled, 3,296 (46.8%) had a viral load done within 12 months of ART initiation. Median time from ART initiation to initial viral load testing was 6.2 (IQR, 5.3-7.3) months. Overall, initial VnS decreased from 9.3% during April 2018 – March 2019 to 5.4% during April 2020 – March 2021 (Figure 5 [a]). There was no significant difference in initial VnS during the COVID-19 period, compared to the pre-COVID-19 period (adjusted Odd Ratio, aOR [95% CI], p-value: 0.79 [95% CI: 0.52–1.20], p=0.264) (Table S4).

**Figure 5.**
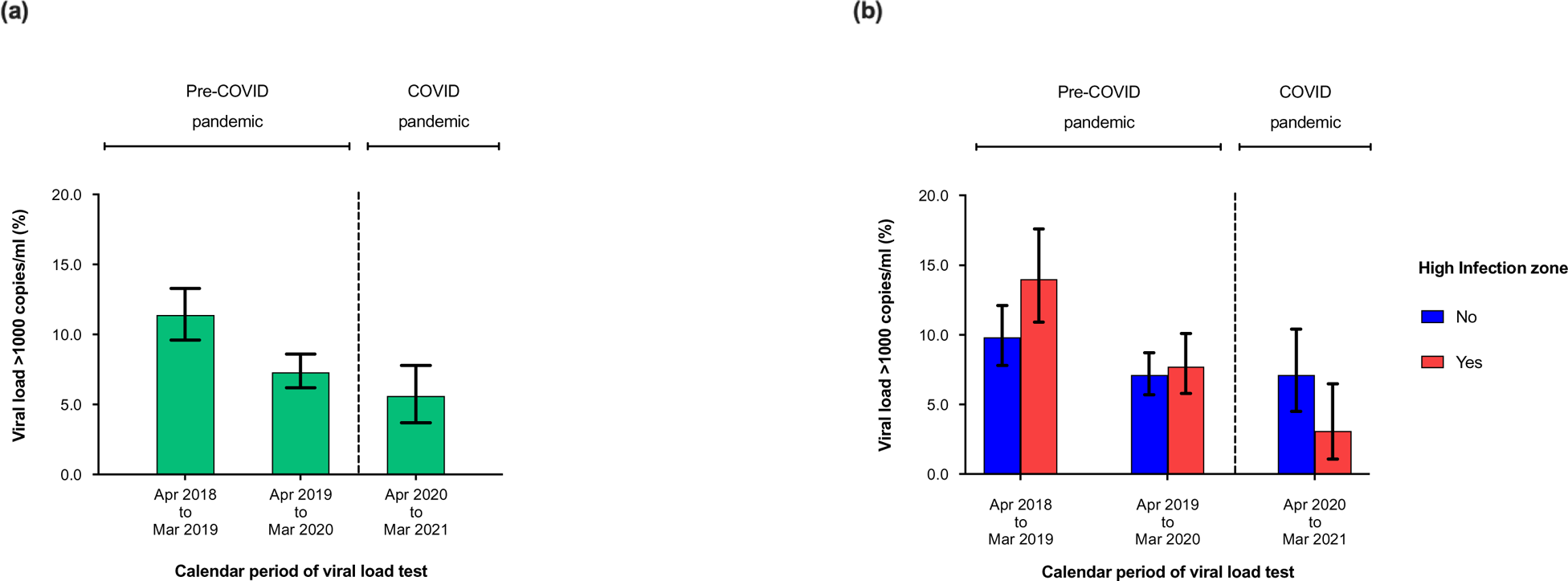
(a) Graph showing initial virologic non-suppression (viral load >1000 copies/ml) after ART initiation by COVID-19 exposure calendar period when compared to the period prior to the pandemic, and (b) by high infection zone counties, amongst HIV infected individuals >15 years using data from the national data warehouse sampling framework in Kenya (April 01^st^ 2018 to March 31^st^ 2021, N=7,046).

From outside the HIZ, initial VnS reduced from 10.1% during April 2018 – March 2019 to 4.2% during April 2020 – March 2021, while that from the HIZ remained relatively stable from 7.9% to 7.6% during the same period (Figure 5 [b]). From outside the HIZ, there was no significant difference in initial VnS during the COVID-19 period, compared to the pre-COVID-19 period (0.66 [0.38–1.16], p=0.137). Similarly, and from the HIZ, there were no significant differences in initial VnS during the COVID-19 period, compared to the pre-COVID-19 period (0.94 [0.53–1.65], p=0.827) (Table 5).

**Table 5.** Effect of the SARS-CoV-2 pandemic, defined as the period after the first documented case when compared to the period prior to the pandemic, on virologic non-suppression amongst HIV infected individuals >15 years starting combination antiretroviral therapy using data from the national data warehouse sampling framework in Kenya (April 01^st^ 2018 to March 31^st^ 2021, N=7,406)*.

| Characteristics |  | High infection zone (No, n=2,102) |  |  |  | High infection zone (Yes, n=1,194) |  |  |  |
| --- | --- | --- | --- | --- | --- | --- | --- | --- | --- |
|  |  | Crude OR<br>(95% CI) | p-value | Adjusted OR<br>(95% CI) | p-value | Crude OR<br>(95% CI) | p-value | Adjusted OR<br>(95% CI) | p-value |
| **Pandemic periods** | Pre-COVID-19 | Ref |  | Ref |  | Ref |  | Ref |  |
|  | COVID-19 | 0.46 (0.27 – 0.77) | 0.004 | 0.66 (0.38 – 1.16) | 0.137 | 0.97 (0.56 – 1.70) | 0.924 | 0.94 (0.53 – 1.65) | 0.827 |
| **Gender** | Female | Ref |  |  |  | Ref |  |  |  |
|  | Male | 0.78 (0.55 – 1.10) | 0.160 | - | - | 1.05 (0.66 – 1.66) | 0.837 | - | - |
| **Age group (years)** | 15.0 – 24.9 | 1.45 (0.85 – 2.48) |  |  |  | 2.99 (1.39 – 6.41) |  | 3.00 (1.40 – 6.44) |  |
|  | 25.0 – 34.9 | 1.30 (0.82 – 2.01) |  |  |  | 1.98 (0.99 – 3.97) |  | 1.98 (0.99 – 3.97) |  |
|  | 35.0 – 44.9 | 1.08 (0.66 – 1.78) |  |  |  | 1.98 (0.96 – 4.09) |  | 1.98 (0.96 – 4.09) |  |
|  | 45.0+ | Ref | 0.449 | - | - | Ref | 0.048 | Ref | 0.034 |
| **First-line ART regimen** | EFV-based | Ref |  | Ref |  | Ref |  |  |  |
|  | DTG-based | 0.40 (0.27 – 0.59) |  | 0.45 (0.29 – 0.68) |  | 0.74 (0.45 – 1.20) |  |  |  |
|  | Others | 0.85 (0.25 – 2.86) |  | 0.87 (0.26 – 2.92) |  | 1.53 (0.43 – 5.37) |  |  |  |
|  | Missing | 0.95 (0.64 – 1.40) | <0.001 | 0.98 (0.66 – 1.44) | 0.001 | 0.75 (0.42 – 1.34) | 0.446 | - | - |
| **Same-day HIV diagnosis and ART start** | No | Ref |  |  |  | Ref |  |  |  |
|  | Yes | 0.79 (0.55 – 1.15) |  |  |  | 0.92 (0.53 – 1.60) |  |  |  |
|  | Missing | 0.93 (0.57 – 1.51) | 0.432 | - | - | 1.52 (0.87 – 2.65) | 0.102 | - | - |
| **ART start to initial viral load (months)** | <3.0 | Ref |  |  |  | Ref |  |  |  |
|  | 3.0 – 5.9 | 1.47 (0.74 – 2.92) |  |  |  | 0.57 (0.29 – 1.11) |  |  |  |
|  | 6.0 – 8.9 | 1.24 (0.63 – 2.46) |  |  |  | 0.78 (0.42 – 1.48) |  |  |  |
|  | 9.0 – 11.9 | 1.26 (0.57 – 2.79) | 0.645 | - | - | 0.37 (0.13 – 1.07) | 0.162 | - | - |

\*HIV viral load test not yet done (n=3,750 [53.2%])

## DISCUSSION

Unintended disruptions from the COVID-19 pandemic threatened to erode gains made in the fight against the HIV epidemic. We aimed to assess impact of the COVID-19 pandemic on HIV care and treatment outcomes amongst individuals starting ART in Kenya. A sampling framework of national routine individual-level service delivery data was randomly sampled. Characteristics of individuals from the sampled population were comparable to that from the sampling framework, and proportional distribution of counties from the sampling framework was reflective of that from the most recent national HIV/AIDS modelling estimates [29], suggesting that our findings maybe generalisable and representative at the national level.

Our findings suggest that same-day HIV diagnosis/ART initiation was on an increase before the COVID-19 pandemic, and that the upward trajectory was sustained despite the pandemic and regardless of whether individuals were from COVID-19 HIZ or not. To our knowledge, there is no literature that has assessed impact of COVID-19 pandemic on time from HIV diagnosis to ART initiation. Data from select health facilities in Nairobi, Kenya, suggest a 51% decline in HIV testing uptake during the COVID-19 period, compared to the pre-COVID-19 period [32]. However, the authors clarify that facility-based HIV testing was already on a steep decline before the COVID-19 period for several reasons including promotion of HIV self-testing, and that the decline stabilized at onset of the COVID-19 pandemic, suggesting that the pandemic did not negatively impact HIV testing.

We observed an overall decline in six-months non-retention over calendar years in Kenya, with the COVID-19 period having significantly lower non-retention compared to the pre-COVID-19 period. The majority (95%) of individuals who had undergone non-retention were LTFU. In a meta-analysis of data to determine outcome of patients LTFU from Africa, the majority (54%) were either known to have died or could not be found (presumed dead) [33]. The decline in non-retention over calendar period in our findings may, therefore, be attributed to a decline in HIV-related mortality. This maybe a reflection of the scale-up of HIV programme interventions including universal test-and-treat, and the more efficacious integrase inhibitor-based first-line regimen. Indeed, our data confirm that during the periods April 2018 – March 2019 and April 2020 – March 2021, same-day HIV diagnosis/ART initiation increased from 63.5% to 79.6%, while that of DTG-based regimen increased from 14.6% to 82.8%, despite the COVID-19 pandemic.

We also observed an overall decline in initial VnS over calendar years, with the most recent (during the COVID-19 period) estimate suggesting 94.6% initial virologic suppression in Kenya. These results suggest that despite the COVID-19 pandemic, Kenya is well on track towards achieving the last milestone in the UNAIDS 95-95-95 targets [22]. Importantly, when compared with the pre-COVID-19 period, there was no significant difference in VnS during the COVID-19 period, suggesting that the pandemic has not yet negatively impacted VnS in Kenya. When compared to estimates from previous national surveys [24, 25], the low level of VnS may also reflect the scale up of more efficacious HIV programme interventions as described above. Early ART initiation [9, 11, 12, 34–36] and the more efficacious DTG-based regimen [37–41] have both been shown to achieve rapid and sustained virologic suppression.

While the COVID-19 pandemic adversely disrupted global health service delivery, our data suggests that impact on HIV care and treatment outcomes in Kenya was less adverse. Within one month of the first reported case of SARS-CoV-2 infection, the Kenyan National AIDS and STI Control Program (NASCOP) had mapped out areas of HIV program concern and prepared a strategic response towards mitigating deleterious effects of the pandemic on HIV service delivery [42]. These included expedited efforts towards stocking up commodities at the county level, promotion and provision of HIV self-testing kits, provision of personal protective equipment to frontline service providers, three- to six-multi-month ART dispensing, promotion of flexible ART delivery models including community groups to distribute ART for decongestion of facilities, use of M-health applications to communicate with clients on continuity of HIV services and enhanced virtual coordination/supervision of HIV service delivery for a sustained response. This timely, strategic and sustained COVID-19 response may have played a critical role in mitigating the adverse effects of the pandemic on HIV care and treatment outcomes and point towards maturity, versatility and resilience of the HIV program in Kenya.

A major strength of our analysis is the use of routine service delivery data collected from health facilities from all over Kenya, suggesting that our findings are a good representation of impact of the COVID-19 pandemic on HIV care and treatment outcomes in the country. However, use of routine service delivery data is not without limitations. First, about 23% and 53% of the study population were missing date of HIV diagnosis and had not received a viral load test since ART initiation, respectively. Unforeseen bias from missing data cannot be ruled out. Second, additional COVID-19 mitigation measures in the HIZ counties were imposed at different times and over different durations. Grouping them may have resulted to a dilution effect in subtle differences between the most adversely affected counties (e.g. Nairobi), over shorter time frames and on the various endpoints. Effect of the COVID-19 pandemic on HIV service delivery at facility- or local-level can therefore not be ruled out, though these were likely transient. Importantly, these differences did not seem to have impacted findings at the national level.

In conclusion, we used nationally representative, routine, individual-level data to assess impact of the COVID-19 pandemic on HIV care and treatment outcomes in Kenya. Overall and when compared to the pre-COVID-19 period, we observed significantly higher levels of same-day HIV diagnosis/ART initiation, lower levels of six-months non-retention and no significant difference with initial VnS during the COVID-19 pandemic period. The higher levels of same-day HIV diagnosis/ART initiation and the lower levels of six-months non-retention were a continuation of a trend that was occurring even before the COVID-19 pandemic. While the COVID-19 pandemic adversely disrupted global health service delivery, our findings suggests that impact on HIV care and treatment outcomes in Kenya was less adverse. Timely, strategic and sustained COVID-19 response by the NASCOP may have played a critical role in mitigating adverse effects of the pandemic on HIV care and treatment services and point towards maturity, versatility and resilience of the HIV program in Kenya. Our findings underscore the value of routine program data for monitoring continuity of HIV care and treatment service delivery and the importance of evidence-based strategies to mitigate the undesired effects of the COVID-19 pandemic in the fight against the HIV epidemic in Kenya.

## Supporting information

Supplementary Information for Covid Manuscript

## Authors Contributions

DOK, VNM, ASH and LN conceived the objectives of the manuscript. DOK, ASH, KJM and FN carried out the data analysis and generated outputs from the case examples. DOK, VNB, and ASH prepared the initial draft of the manuscript. LN, LO, AO, MMN and GO critically reviewed the initial draft and provided insightful feedback. All authors reviewed, proof-read and approved the final manuscript.

## Attribution of support

This analysis is based on data from the national data warehouse (NDW) which is supported by the President’s Emergency Plan for AIDS Relief (PEPFAR) through the Centers for Disease Control and Prevention (CDC) under the terms of mechanism ID 18214. At the time of the analysis, the NDW was under the technical management of the Palladium group with oversight from the National HIV/AIDS Strategic Information and Evaluation technical working group. Views expressed in this publication are those of the authors and not necessarily those of PEPFAR, CDC or the Palladium group.

## Acknowledgement

We acknowledge the contribution of all HIV-infected individuals and health care providers from all the facilities that have uploaded data to the NDW. We are thankful to health facility administrators, service delivery partners, donor agencies and other stakeholders for their continued support with the NDW project. We are particularly grateful to the ministry of health at both national and county level for the oversight and leadership on the NDW project.

## Data availability

De-identified data used in this analysis is available from the National EMR data warehouse in Kenya managed by the Ministry of Health.

