## Supplementary Information for Covid Manuscript for "IMPACT OF THE COVID-19 PANDEMIC ON ROUTINE HIV CARE AND ANTIRETROVIRAL TREATMENT OUTCOMES IN KENYA: A NATIONALLY REPRESENTATIVE ANALYSIS"

**Author Affiliations**: (1) Division for Global HIV & TB (DGHT), Center for Global Health, US Centres for Disease Control and Prevention (CDC), Nairobi, Kenya; (2) National AIDS and STI Control Program (NASCOP), Ministry of Health, Nairobi, Kenya; (3) KEMRI/Wellcome Trust Research Programme, Kilifi, Kenya; (4) Epidemiology and Data Use, Palladium Group, Nairobi, Kenya; (5) Health Population and Nutrition, United States Agency for International Development (USAID), Nairobi, Kenya; and, (6) Strategic Information, Military HIV Research Program/Walter Reed Army Institute of Research (MHRP/WRAIR), Nairobi, Kenya.

**^§^Corresponding Author:**

Dr. Davies O. Kimanga

P. O. Box 606 - 00621 Nairobi, Kenya

**Key words:** HIV, COVID-19, antiretroviral therapy, test-and-treat, retention, virologic suppression.

**SUPPLEMENTARY TABLES**

**Supplementary Table 1.** Distribution of HIV infected individuals >15 years old, starting combination antiretroviral therapy, included in the national data warehouse sampling framework and randomly sampled, by COVID-19 exposure status in Kenya (April 2018 to March 2021, N=7,046).

| Characteristics | | Apr 2018  To  Mar 2019  (n=2,505) | Apr 2019  To  Mar 2020  (n=2,538) | Apr 2020  To  Mar 2021  (n=2,003) | Overall  (n=7,046) |
| --- | --- | --- | --- | --- | --- |
| Gender | Female  Male | 1,681 (67.1)  824 (32.9) | 1,692 (66.7)  846 (33.3) | 1,330 (66.4)  673 (33.6) | 4,703 (66.7)  2,343 (33.3) |
| Age group (years) | 15.0 – 24.9  25.0 – 34.9  35.0 – 44.9  45.0 – 54.9  55.0+ | 430 (17.2)  909 (36.3)  652 (26.0)  337 (13.5)  177 (7.1) | 434 (17.1)  931 (36.7)  666 (26.2)  343 (13.5)  164 (6.5) | 344 (17.2)  718 (35.9)  577 (28.8)  249 (12.4)  115 (5.7) | 1,208 (17.1)  2,558 (36.3)  1,895 (26.9)  929 (13.2)  456 (6.5) |
| First-line ART regimen | NVP-based  EFV-based  DTG-based  Others  Missing | 51 (2.0)  1,375 (54.9)  365 (14.6)  34 (1.4)  680 (27.2) | 3 (0.1)  863 (34.0)  1,088 (42.9)  20 (0.8)  564 (22.2) | 1 (0.1)  121 (6.0)  1,658 (82.8)  7 (0.4)  216 (10.8 | 55 (0.8)  2,359 (33.5)  3,111 (44.2)  61 (0.9)  1,460 (20.7) |
| HIV diagnosis to ART initiation (days) | Same day  1-14 days  15-90 days  91+ days  Missing | 1,235 (49.3)  212 (8.5)  165 (6.6)  333 (13.3)  560 (22.4) | 1,308 (51.5)  189 (7.5)  128 (5.0)  197 (7.8)  716 (28.2) | 1,294 (64.6)  149 (7.4)  86 (4.3)  97 (4.8)  377 (18.8) | 3,837 (54.5)  550 (7.8)  379 (5.4)  627 (8.9)  1,653 (23.5) |
| ART start to initial viral load (months) | <3.0  3.0 – 5.9  6.0 – 8.9  9.0 – 11.9  <Missing | 121 (4.8)  459 (18.3)  589 (23.5)  144 (5.8)  1,192 (47.6) | 100 (3.9)  464 (18.3)  635 (25.0)  187 (7.4)  1,152 (43.4) | 71 (3.5)  253 (12.6)  240 (12.0)  33 (1.7)  1,406 (70.2) | 292 (4.1)  1,176 (16.7)  1,464 (20.8)  364 (5.2)  3,750 (53.2) |

**Supplementary Table 2.** Overall effect of the COVID-19 pandemic, defined as period after the first documented case when compared to the period prior to the pandemic, on time from a HIV diagnosis to combination antiretroviral therapy start (Same day ART initiation) amongst HIV infected individuals aged >15 years using data from the national data warehouse sampling framework in Kenya (April 2018 to March 2021, N=7,046)*.

| Characteristics |  | Same day, n (%) | Crude RR (95% CI) | p-value | Adjusted RR (95% CI) | p-value |
| --- | --- | --- | --- | --- | --- | --- |
| Pandemic periods | Pre-COVID-19  COVID-19 | 2,543 (67.5)  1,294 (79.6) | Ref  1.16 (1.12 – 1.20) | <0.001 | Ref  1.09 (1.04 – 1.13) | <0.001 |
| Gender | Female  Male | 2,599 (71.8)  1,238 (69.8) | Ref  0.97 (0.93 – 1.00) | 0.065 | Ref  0.97 (0.93 – 1.00) | 0.065 |
| Age group (years) | 15.0 – 24.9  25.0 – 34.9  35.0 – 44.9  45.0 – 54.9  55.0+ | 759 (78.8)  1,435 (72.3)  985 (69.1)  437 (64.1)  221 (65.4) | 1.18 (1.09 – 1.28)  1.10 (1.02 – 1.19)  1.06 (0.98 – 1.14)  1.00 (0.91 – 1.09)  Ref | <0.001 | 1.15 (1.07 – 1.25)  1.08 (1.01 – 1.17)  1.04 (0.97 – 1.13)  0.98 (0.90 – 1.07)  Ref | <0.001 |
| First-line ART regimen | EFV-based  DTG-based  Others  Missing | 1,184 (69.4)  1,865 (76.7)  6 (11.3)  782 (65.0) | Ref  1.09 (1.05 – 1.13)  0.18 (0.07 – 0.47)  0.92 (0.88 – 0.96) | <0.001 | Ref  1.07 (1.02 – 1.13)  0.18 (0.07 – 0.49)  0.93 (0.89 – 0.98) | <0.001 |

*Missing date of HIV diagnosis (n=1,653 [23.5%])

**Supplementary Table 3.** Overall effect of the COVID-19 pandemic, defined as period after the first documented case when compared to the period prior to the pandemic, on attrition from combination antiretroviral therapy start amongst HIV infected individuals aged >15 years using data from the national data warehouse sampling framework in Kenya (April 2018 to March 2021, N=7,046)*.

| Characteristics |  | Attrition, d/Y  (rate per 100 pyo) | Crude HR  (95% CI) | p-value | Adjusted HR  (95% CI) | p-value |
| --- | --- | --- | --- | --- | --- | --- |
| Pandemic periods | Pre-COVID-19  COVID-19 | 1,448/149.5 (9.69)  394/90.8 (4.34) | Ref  0.54 (0.49 – 0.61) | <0.001 | Ref  0.66 (0.58 – 0.74) | <0.001 |
| Gender | Female  Male | 1190/161.9 (7.35)  652/78.5 (8.31) | Ref  1.12 (1.02 – 1.23) | 0.019 | Ref  1.19 (1.07 – 1.32) | 0.001 |
| Age group (years) | 15.0 – 24.9  25.0 – 34.9  35.0 – 44.9  45.0 – 54.9  55.0+ | 334/38.9 (8.58)  681/88.6 (7.69)  482/64.5 (7.48)  220/32.5 (6.77)  125/15.9 (7.87) | 1.08 (0.88 – 1.33)  1.01 (0.83 – 1.22)  0.96 (0.79 – 1.18)  0.86 (0.69 – 1.07)  Ref | 0.115 | - |  |
| First-line ART regimen | EFV-based  DTG-based  Others  Missing | 600/79.5 (7.55)  610/115.3 (5.29)  34/3.5 (9.75)  598/42.0 (14.23) | Ref  0.72 (0.64 – 0.81)  1.13 (0.80 – 1.59)  1.91 (1.70 – 2.14) | <0.001 | Ref  0.82 (0.72 – 0.94)  1.16 (0.82 – 1.64)  1.98 (1.76 – 2.23) | <0.001 |
| Same day HIV diagnosis and ART start | No  Yes  Missing | 360/55.6 (6.47)  1000/133.3 (7.50)  482/51.4 (9.38) | Ref  1.16 (1.03 – 1.31)  1.33 (1.15 – 1.53) | <0.001 | Ref  1.35 (1.19 – 1.52)  1.50 (1.30 – 1.73) | <0.001 |

**Supplementary Table 4.** Overall effect of the COVID-19 pandemic, defined as period after the first documented case when compared to the period prior to the pandemic, on initial virologic non-suppression (VnS) amongst HIV infected individuals aged >15 years using data from the national data warehouse sampling framework in Kenya (April 2018 to March 2021, N=7,046)*.

| Characteristics |  | VnS, n (%) | Crude OR (95% CI) | p-value | Adjusted OR (95% CI) | p-value |
| --- | --- | --- | --- | --- | --- | --- |
| Pandemic periods | Pre-COVID-19  COVID-19 | 225/2,699 (8.3)  32/597 (5.4) | Ref  0.62 (0.43 – 0.91) | 0.015 | Ref  0.79 (0.52 – 1.20) | 0.264 |
| Gender | Female  Male | 182/2,241 (8.1)  75/1,055 (7.1) | Ref  0.87 (0.65 – 1.14) | 0.312 | - | - |
| Age group (years) | 15.0 – 24.9  25.0 – 34.9  35.0 – 44.9  45.0 – 54.9  55.0+ | 50 (10.1)  101 (8.3)  67 (7.4)  31 (6.7)  8 (3.7) | 2.96 (1.38 – 6.35)  2.37 (1.14 – 4.94)  2.11 (1.00 – 4.46)  1.87 (0.85 – 4.14)  Ref | 0.042 | 2.54 (1.18 – 5.20)  2.10 (1.00 – 4.41)  1.97 (0.93 – 4.19)  1.87 (0.84 – 4.15)  Ref | 0.134 |
| First-line ART regimen | EFV-based  DTG-based  Others  Missing | 118/1,190 (9.9)  71/1,348 (5.3)  6/56 (10.7)  62/702 (8.8) | Ref  0.51 (0.37 – 0.69)  1.09 (0.46 – 2.60)  0.88 (0.64 – 1.22) | <0.001 | Ref  0.58 (0.42 – 0.82)  1.16 (0.48 – 2.77)  0.93 (0.67 – 1.29) | 0.010 |
| Same day HIV diagnosis and ART start | No  Yes  Missing | 68/828 (8.2)  124/1,772 (7.0)  65/696 (9.3) | Ref  0.84 (0.62 – 1.14)  1.15 (0.81 – 1.64) | 0.132 | - | - |
| Duration from ART start to initial viral load test (months) | <3.0  3.0 – 5.9  6.0 – 8.9  9.0 – 11.9 | 24/292 (8.2)  93/1,176 (7.9)  116/1,464 (7.9)  24/364 (6.6) | Ref  0.96 (0.60 – 1.53)  0.96 (0.61 – 1.52)  0.79 (0.44 – 1.42) | 0.836 | - | - |

*HIV viral load test not yet done (n=3,750 [53.2%])
